# Mental healthcare utilisation and costs before and after dementia diagnosis: evidence from electronic health records

**DOI:** 10.64898/2026.06.02.26354695

**Authors:** Gillian Eaglestone, Charlotte Stoner, Rosana Pacella, Paul McCrone

## Abstract

**Objectives:** To describe secondary mental healthcare utilisation and associated costs among patients diagnosed with dementia or mild cognitive impairment (MCI).

**Design:** Retrospective cohort study using routinely collected electronic health record data.

**Setting:** Secondary mental healthcare services within a large NHS mental health provider in South London, UK.

**Participants:** Adults aged 18 years or older with a recorded diagnosis of dementia or MCI between 1 January 2010 and 31 December 2020. Patients surviving less than one year after diagnosis were excluded. The final cohort comprised 16,081 individuals.

**Primary and secondary outcome measures:** Service utilisation and NHS mental health service costs during the 12 months before and after diagnosis, including inpatient, outpatient and memory clinic contacts.

**Results:** The proportion of patients with at least one recorded mental health service contact declined from 91% in the 12 months before diagnosis to 69% after diagnosis. Among service users, mean NHS mental health costs increased from £1,497 to £2,177 per person following diagnosis (mean increase £680; p<0.001), driven primarily by inpatient care. Dementia diagnosis, younger age, male gender, living alone, greater cognitive impairment and higher clinical symptom burden were independently associated with higher costs. Ethnic differences in service use and costs were also observed.

**Conclusions:** Although overall service engagement declined following diagnosis, costs increased among those continuing to access care, indicating greater intensity of service use. Understanding patterns of secondary mental healthcare utilisation and associated costs may help inform planning and resource allocation within dementia services.

**Strengths and limitations of this study:**

- This study used a large retrospective cohort of 16,081 adults with mild cognitive impairment or dementia identified from routinely collected electronic health records within a major UK mental health provider.
- The Clinical Record Interactive Search (CRIS) platform enabled analysis of service utilisation and costs across inpatient, outpatient, and memory clinic services before and after diagnosis.
- The study examined healthcare utilisation within the same individuals during the 12 months before and after diagnosis, allowing comparison of changes over time.
- The analysis was restricted to secondary mental healthcare contacts recorded within South London and Maudsley NHS Foundation Trust and did not capture primary care, acute hospital care, social care, medication costs, or informal care.
- Routinely collected electronic health record data may contain missing or incomplete information, particularly for variables such as living situation and care home residence.

## Introduction

Dementia is associated with high healthcare use and costs (1). People living with dementia (PLwD) or mild cognitive impairment (MCI) have higher rates of hospital admission, re-admission, emergency admission, and longer stays than those without cognitive impairment (2,3). Hospitalisation is linked to adverse outcomes, including functional decline, complications, and higher mortality (4). Risk of admission is influenced by comorbidities, polypharmacy, falls history, and caregiver burden (5,6). PLwD frequently experience comorbid mental health, cardiovascular, and metabolic conditions, which complicate care and increase hospital use (7). A substantial proportion of service use is driven by crises in community care, which may be preventable (8). Supporting PLwD in the community can reduce reliance on hospital or institutional care and may be more cost-effective.

Although the economic burden of dementia has been widely documented, much of the literature has focused on overall healthcare, social care, or societal costs. Less is known about patterns of secondary mental healthcare utilisation surrounding the point of diagnosis and how demographic, clinical, and social characteristics influence service costs within specialist mental health services. Understanding these patterns may help identify groups with greater service needs and inform planning of dementia care pathways within secondary mental healthcare settings.

Using routinely collected data from the Clinical Record Interactive Search (CRIS) platform, this study examined patterns of specialist mental healthcare utilisation and associated costs in the 12 months before and after a recorded diagnosis of MCI or dementia within a large, ethnically diverse urban population. We also explored demographic, clinical, and social factors associated with variation in service costs.

### Aim

This study examined mental health service use and associated NHS costs in the year before and after a diagnosis of dementia or MCI, using a retrospective cohort of patients referred to mental health services within a UK NHS Trust. The aim was to quantify changes in service utilisation and costs surrounding diagnosis and to identify factors associated with cost variation.

## Methods

We conducted a study of mental health service use and costs using a secondary dataset from a UK cohort of patients diagnosed with MCI or dementia and registered with South London and Maudsley NHS Foundation Trust (SLaM). Clinical records from SLaM’s electronic health record system are linked to the Clinical Record Interactive Search (CRIS) database, which provides secure, anonymised access to routinely collected electronic health record data for approved research.

The CRIS database captures contacts recorded within SLaM secondary mental healthcare services. Consequently, the service utilisation data used in this study reflect specialist mental health service contacts, including memory clinic, outpatient, and inpatient services recorded within SLaM. The dataset does not capture healthcare use occurring outside SLaM services, including primary care consultations, acute hospital care, social care services, informal care, or medication costs. Therefore, the costs reported represent secondary mental healthcare costs rather than the total healthcare or societal costs associated with dementia.

CRIS contains de-identified clinical data not originally collected for research purposes. Patients may opt out of inclusion in CRIS, ensuring protection of autonomy while enabling secondary analysis for service evaluation. Ethical approval for the use of anonymised electronic health records from the CRIS database was granted by the South-Central Oxford C Research Ethics Committee (Reference: 23/SC/0257). Individual informed consent was not required because the study used de-identified routinely collected data accessed under CRIS governance procedures. All analyses were conducted within secure governance frameworks in accordance with NHS data protection standards.

SLaM provides specialist mental health and dementia services locally and nationally, with a catchment area of 1.3 million people across the London boroughs of Croydon, Lambeth, Lewisham, and Southwark (SLaM NHS Foundation Trust, 2024). Southeast London is ethnically diverse (SLaM NHS Foundation Trust, 2024) and has high levels of deprivation, health inequalities, and physical and mental health burden (10).

### Study participants

Records of all patients aged 18 years or older with a primary diagnosis of MCI or dementia between 01/01/2010 and 31/12/2020 were retrieved. Dementia diagnosis was derived using International Classification of Disease (ICD-10) (World Health Organization, 1993). Patients that survived less than one year after diagnosis were excluded, to allow sufficient post-diagnosis service use analysis. Patients living in nursing and care homes were included. Individuals with MCI were included because MCI is commonly assessed and managed within the same specialist memory service pathways as dementia. Including both groups allowed examination of service utilisation and costs across the spectrum of cognitive impairment encountered within secondary mental healthcare services.

### Cohort characteristics

Key demographic and clinical variables were extracted, including gender, age, ethnicity, living situation, primary diagnosis, Mini-Mental State Examination (MMSE) scores, and Health of the Nation Outcome Scales (HoNOS) scores. For both MMSE and HoNOS, the earliest and most recent scores recorded within the study period were used to assess changes in cognitive and clinical status over time. Cognitive impairment was characterised using recorded MMSE scores (12). For descriptive and analytical purposes, MMSE scores were grouped into categories reflecting levels of cognitive impairment (≥27, 21-26, 10-20, ≤9). These categories were used as pragmatic indicators of cognitive status within the dataset and should not be interpreted as comprehensive measures of dementia severity. For descriptive and analytical purposes, HoNOS scores were also categorised into levels of clinical symptom burden.

Descriptive statistics were used to summarise cohort characteristics. Categorical variables are presented as frequencies and percentages, and continuous variables as means and standard deviations.

### Statistical Analysis and Costing

Univariable analyses were conducted to examine patterns of service use and costs across predefined demographic and clinical subgroups, including gender, primary diagnosis, ethnicity, living situation, age, MMSE category, and HoNOS category. Age was grouped for descriptive analyses to support interpretation but treated as a continuous variable in regression models to retain statistical power.

All analyses compared service use and costs across two periods: 12 months before and the 12 months after diagnosis. Outcomes included type and number of contacts (inpatient admissions, outpatient contacts, and memory clinic appointments) and total NHS mental health service costs per person. Service use was identified using unique patient identifiers. Only attended appointments were included; missed appointments and contacts occurring during inpatient admissions were excluded. Frequencies, percentages, means, and standard deviations were used to summarise service utilisation, and costs were reported as means and standard deviations.

Analyses were conducted for the full cohort and for “service users,” defined as individuals with at least one recorded service contact during the study period. Total NHS mental health service costs were calculated using standardised UK unit cost references from the Personal Social Services Research Unit (PSSRU) Unit Costs of Health and Social Care 2022 and the NHS National Cost Collection 2021/22 (13,14). Service categories included inpatient admissions, outpatient contacts, and memory clinic appointments. Inpatient services comprised admissions to SLaM mental health inpatient wards. Outpatient services included contacts with specialist mental health services outside memory clinics, including community and clinic-based appointments. Memory clinic services comprised contacts recorded within specialist memory assessment and diagnostic services. Categories included both direct (in-person) and indirect (telephone) contacts where recorded. All costs are presented in 2024 UK pounds sterling.

Differences in service contacts and mean total costs between pre- and post-diagnosis periods were tested using paired t-tests. Distributional assumptions were assessed using histograms and the Shapiro–Wilk test. Multivariable linear regression models were used to explore demographic and clinical predictors of total service costs, with coefficients reported alongside 95% confidence intervals; p-values < 0.05 were considered statistically significant. Data cleaning was undertaken in Microsoft Excel, and analyses were completed using Stata version 15.1 (15).

### Missing and incomplete data

Some variables contained substantial missing data, most notably living situation, which was inconsistently recorded in the electronic health record system. HoNOS scores had been pre-adjusted within the dataset to account for incomplete assessments, with around one-third of cases classified as ‘unknown’. All analyses were conducted using available data, and missing categories were retained and reported where relevant.

### Sensitivity analysis

One-way sensitivity analyses were conducted to assess the robustness of cost estimates by varying unit costs assumptions. Baseline cost values were increased by 25% and 50% to examine the impact of variation in service pricing on overall findings.

## Results

### Cohort characteristics

The cohort consisted of 16,081 adults with MCI or dementia (Table 1). The majority were female (61%), and most were aged 65 years or older at diagnosis (94%). Dementia accounted for 82% of recorded diagnoses. MCI accounted for the remaining 18% of the cohort and was retained in analyses because MCI and dementia are commonly assessed and managed within the same specialist memory service pathways. Two-thirds (67%) identified as White ethnicity, and 38% were living with someone at the time of assessment. Mean MMSE scores were lower at post-diagnosis assessment than at pre-diagnosis assessment, indicating greater cognitive impairment among those assessed after diagnosis, while HoNOS scores increased over the same period, suggesting greater clinical symptom burden.

**Table 1.** Cohort characteristics – table to be placed here.

| Characteristic | N (%) or Mean (SD) |
| --- | --- |
| **Gender** |  |
| Female | 9,760 (61%) |
| Male | 6,321 (39%) |
| **Age** | 79 (9.51) |
| 18-64 years | 1,105 (7%) |
| 65-84 years | 9,909 (62%) |
| ≥85 years | 5,067 (32%) |
| **Primary diagnosis** |  |
| Dementia | 13,229 (82%) |
| MCI | 2,852 (18%) |
| **Ethnicity** |  |
| White British | 9,002 (56%) |
| White non-British | 1,696 (11%) |
| Black | 2,979 (19%) |
| Mixed race | 151 (1%) |
| Asian | 1,027 (6%) |
| 'Other' ethnicity | 414 (3%) |
| Unknown | 812 (5%) |
| **Living situation** |  |
| Lived alone | 4,661 (29%) |
| Lived with someone | 6,162 (38%) |
| Unknown | 5,258 (33%) |
| **MMSE baseline** | 19.81 (6.32) |
| **HoNOS baseline** | 9.75 (5.43) |

*HoNOS Health of the Nation Outcome Scales, MMSE Mini-Mental State Examination*

### Comparison of service use and costs

The proportion of patients with at least one recorded contact across any service declined from 91% in the 12 months pre-diagnosis to 69% post-diagnosis. Patterns of utilisation differed by service type (Table 2). Among in-person outpatient users, mean contacts increased from 3.25 to 5.44 (p < 0.001), indicating greater intensity of use among those who remained engaged. Inpatient admissions were less common overall (2% pre-diagnosis; 3% post-diagnosis) but mean number of inpatient days per admitted patient increased from 101 to 127 days. Memory clinic attendance declined in proportion, with relatively stable mean contacts per user.

**Table 2.** Comparison of service use (pre- and post-diagnosis) – to be placed here.

|  | Pre-diagnosis |  |  |  | Post-diagnosis |  |  |  |
| --- | --- | --- | --- | --- | --- | --- | --- | --- |
|  | Service users (n, %) | Service user contacts (mean, SD) | Service user costs (£ mean, SD) | Whole cohort costs £ (mean, SD) | Service users (n, %) | Service user contacts (mean, SD) | Service user costs £ (mean, SD) | Whole cohort costs £ (mean, SD) |
| **Inpatient services** |  |  |  |  |  |  |  |  |
| Inpatient days | 346 (2) | 101.34 (118.22) | 34,557 (40,312) | 744 (7,747) | 462 (3) | 127.50 (111.89) | 43,477 (38,156) | 1,249 (9,720) |
| **Outpatient services** |  |  |  |  |  |  |  |  |
| In-person contacts | 14,446 (90) | 3.25 (5.59) | 764 (1,313) | 686 (1,266) | 10,980 (68) | 5.44 (8.93) | 1,278 (2,098) | 872 (1,833) |
| Indirect contacts | 159 (1) | 1.28 (0.58) | 75 (34) | 1 (8) | 348 (2) | 3.58 (4.31) | 210 (253) | 5 (48) |
| **Memory clinics** |  |  |  |  |  |  |  |  |
| In-person memory clinic contacts | 8,876 (55) | 1.83 (1.18) | 120 (78) | 67 (83) | 6,212 (39) | 1.98 (1.46) | 131 (96) | 51 (87) |
| Indirect memory clinic contacts | 135 (1) | 1.24 (0.51) | 21 (8) | <0 (2) | 176 (1) | 2.18 (2.03) | 36 (33) | <0 (5) |

Outpatient services exclude memory clinic contacts. Memory clinic contacts relate to specialist cognitive assessment and memory services recorded within SLAM.

Analysis of service use and associated costs demonstrated changes in both utilisation patterns and expenditure following diagnosis. Inpatient care remained the costliest service in both periods. The proportion of patients admitted increased slightly from 2% to 3%, and the mean number of inpatient days per admitted patient rose from 101.34 to 127.50 days.

Fewer patients accessed in-person outpatient services after diagnosis (68% vs 90% pre-diagnosis); however, among those who did attend, the mean number of contacts increased from 3.25 to 5.44, suggesting more intensive use of services per patient.

Memory clinic attendance was more common before diagnosis (55% vs 39% for in-person contacts). Among those attending, the mean number of contacts increased modestly (1.83 to 1.98). Indirect memory clinic contacts remained rare in both periods (1%), but the mean number of contacts among those who received them increased from 1.24 to 2.18.

Changes in utilisation aligned with shifts in costs. Inpatient care accounted for the largest share of expenditure in both periods, with mean inpatient costs per admitted patient rising from £34,557 to £43,477. At the whole-cohort level, inpatient costs increased from £744 to £1,249 per person.

For in-person outpatient services, mean costs per service user increased from £764 to £1,278 following diagnosis. Despite fewer patients accessing outpatient services, whole-cohort outpatient costs increased from £686 to £872 per person, reflecting higher use among those who did attend.

Memory clinic costs per service user increased modestly (from £120 to £131), but whole-cohort costs declined (£67 to £51), consistent with reduced overall utilisation post-diagnosis. Indirect contacts contributed minimally to total costs in both periods.

### Comparison of service use and costs by subgroup

Service use and costs varied by age, primary diagnosis, gender and clinical symptom burden, and other demographic factors (S1–S7). Younger patients (under 65 years) consistently incurred the highest costs, particularly for inpatient care, while those aged 85 years and over had the lowest costs across most service categories.

Patients with dementia had substantially higher service use and associated costs than those with MCI, especially for inpatient and outpatient care. Although individuals with MCI accessed memory clinics more frequently before diagnosis, overall costs remained higher among those with dementia.

Greater clinical symptom burden, as measured by HoNOS, was strongly associated with increased post-diagnosis costs. Patients with high HoNOS scores had substantially higher inpatient costs than those with lower scores, while memory clinic use declined as clinical symptom burden increased. A similar pattern was observed for cognitive impairment: inpatient costs increased with greater cognitive impairment, whereas memory clinic use declined among patients with greater cognitive impairment.

Gender differences were most evident in inpatient care, with males experiencing higher admission rates and greater inpatient costs than females. Ethnic differences in service use were also observed, with Black patients showing higher inpatient costs post-diagnosis compared with other groups. Patients living alone had longer inpatient stays and incurred higher inpatient costs than those living with someone.

Interpretation of living-situation findings was limited by missing data, as approximately one-third of the cohort did not have this information recorded.

### Comparison of total costs

Total NHS mental health service costs increased substantially following diagnosis, with considerable variation between individuals, largely driven by a small number of patients with very high service use (Table 3). Across all demographic and clinical subgroups, post-diagnosis costs were consistently higher than pre-diagnosis costs (S8).

**Table 3.**
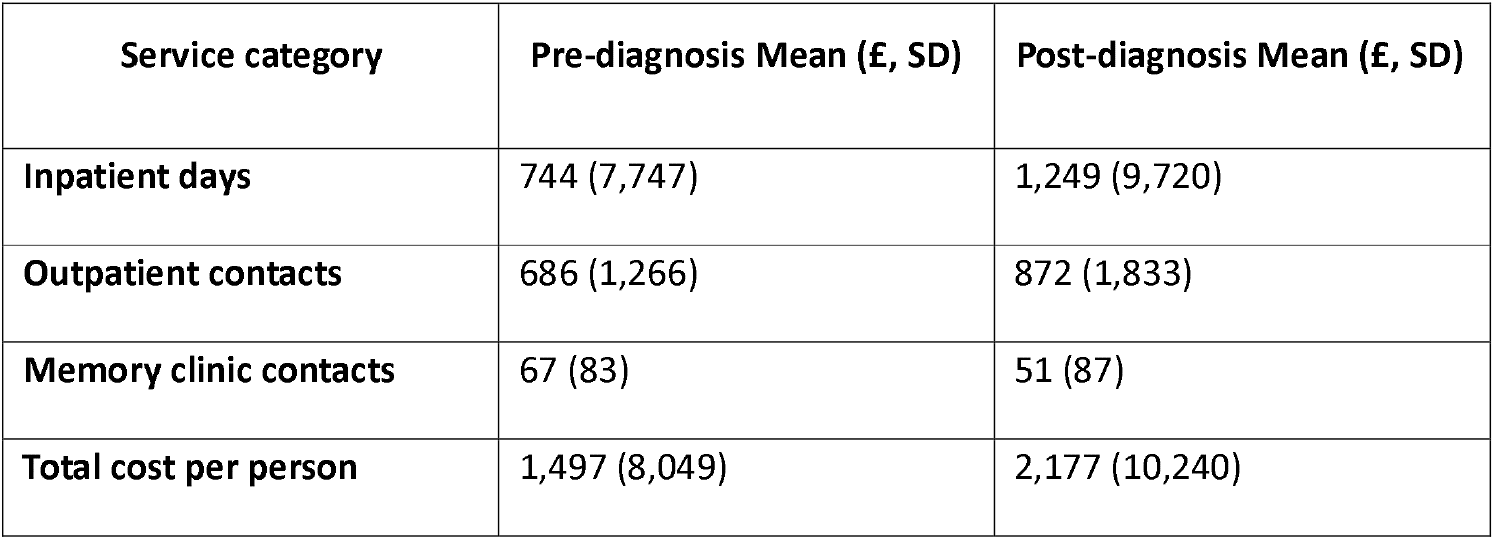
Mean Total Costs per Person – to be placed here.

| Service category | Pre-diagnosis Mean (£, SD) | Post-diagnosis Mean (£, SD) |
| --- | --- | --- |
| **Inpatient days** | 744 (7,747) | 1,249 (9,720) |
| **Outpatient contacts** | 686 (1,266) | 872 (1,833) |
| **Memory clinic contacts** | 67 (83) | 51 (87) |
| **Total cost per person** | 1,497 (8,049) | 2,177 (10,240) |

Although a smaller proportion of patients accessed services following diagnosis, mean total NHS mental health costs increased from £1,497 to £2,177 per person, reflecting greater intensity of service use among those who remained engaged with care.

Patients with greater cognitive impairment and higher clinical symptom burden had the highest total costs, particularly for inpatient care. For example, those with greater cognitive impairment incurred mean total costs of £2,337 pre-diagnosis and £3,853 post-diagnosis, while patients with HoNOS scores ≥25 had the highest overall costs (£7,762 and £17,084, respectively).

Younger patients and males also incurred higher post-diagnosis costs. Patients under 65 years had the highest mean total costs (£2,272 pre-diagnosis; £3,049 post-diagnosis), with costs declining steadily with increasing age. Males had higher post-diagnosis costs than females (£2,644 vs £1,874). Patients diagnosed with dementia had substantially higher costs than those diagnosed with MCI (£2,434 vs £986 post-diagnosis), reflecting more complex clinical needs.

Ethnic differences in total costs were observed. Black patients had the highest post-diagnosis inpatient costs (£1,788), whereas Asian patients had lower inpatient costs (£479). Patients living alone experienced longer inpatient admissions and higher associated costs than those living with someone (£1,398 vs £703 post-diagnosis), although interpretation is limited by the large proportion of missing data on living status.

### Inferential analysis

Total cost data were positively skewed, and distributional assumptions were assessed using histograms and the Shapiro–Wilk test. Given the large sample size, paired t-tests were used to compare mean costs in the 12 months before and after diagnosis. Among service users, mean total NHS mental health costs increased from £1,497 pre-diagnosis to £2,177 post-diagnosis, a mean increase of £680 per person (p < 0.001).

Subgroup analyses showed statistically significant differences across age, diagnosis, ethnicity, cognitive impairment, and clinical symptom burden categories (all p < 0.05), with the strongest effects associated with clinical symptom burden.

Multivariable linear regression identified dementia diagnosis, male gender, younger age, living alone, greater cognitive impairment and higher clinical symptom burden as independent predictors of increased total costs (Table 4).

**Table 4.** Multivariable regression: effects on total service costs pre-/post-diagnosis – to be placed here.

|  | Multivariable regression |  |  |  |
| --- | --- | --- | --- | --- |
|  | Pre-diagnosis |  | Post-diagnosis |  |
| Independent variable | Coef. | 95% CI | Coef. | 95% CI |
| **Gender (male)** | 493 | 234 to 752 | 1,493 | 753 to 2,233 |
| **Age** | -61 | -77 to -45 | -179 | -225 to -133 |
| **Primary diagnosis (MCI)** | -707 | -1,061 to -352 | -1,108 | -2,154 to -63 |
| **Ethnicity** |  |  |  |  |
| White non-British | 451 | 33 to 870 | -645 | -1,788 to 498 |
| Black | 88 | -2,478 to 423 | 481 | -462 to 1,423 |
| Mixed | 269 | -1,077 to 1,614 | -1,010 | -4,766 to 2,747 |
| Asian | -78 | -603 to 446 | -1,740 | -3,292 to -188 |
| 'Other' ethnicity | -313 | -1,126 to 499 | -1,761 | -4,426 to -905 |
| Unknown | -582 | -1,246 to 83 | -1,342 | -4,006 to 1,322 |
| **Living situation** |  |  |  |  |
| Living with someone | -361 | -702 to -19 | -1,317 | -2,131 to -503 |
| Unknown | -165 | -488 to 158 | 651 | -346 to 1,647 |
| **MMSE category** | 25 | 3 to 47 | -40 | -102 to 22 |
| **HoNOS category** | 188 | 163 to 214 | 517 | 444 to 591 |
| **Constant** | 4,116 | 2,641 to 5,590 | 14,347 | 10,098 to 18,596 |

HoNOS - Health of the Nation Outcome Scales, MMSE – Mini-Mental State Examination, MCI – mild cognitive impairment
MMSE and HoNOS were entered into regression models as categorical variables representing levels of cognitive impairment and clinical symptom burden.

Sensitivity analyses, in which unit costs were increased by 25% and 50%, did not change the direction or interpretation of findings, supporting the robustness of the results (S9).

## Discussion

This analysis of a UK cohort of patients with dementia or MCI receiving NHS mental health services identified distinct patterns in service use and cost drivers. Only 3% of patients experienced an inpatient admission, substantially lower than rates reported in previous studies (16,17). This may partly reflect the impact of the COVID-19 pandemic, during which non-emergency admissions were reduced due to wider service pressures within the NHS (18).

Inpatient care was the costliest service category and costs increased post-diagnosis, particularly among patients with dementia. Living alone was associated with higher inpatient costs, consistent with research linking reduced informal support to increased hospital utilisation. Reduced availability of community services during the pandemic may also have intensified reliance on acute care.

Men incurred higher inpatient and overall service costs than women. Previous studies have reported gender differences in healthcare utilisation among people with dementia, although the mechanisms underlying these differences remain unclear. The current dataset did not allow exploration of the factors contributing to higher costs among men. Age showed an inverse association with total NHS costs, with younger patients (<65 years) incurring higher expenditure than older patients. This pattern may reflect differences in care pathways: younger individuals may receive more intensive specialist input, while older individuals with advanced disease may transition toward social care settings not captured within NHS mental health cost data.

Differences in service utilisation were observed across ethnic groups. Previous research has suggested that factors such as cultural and language barriers may contribute to variation in access to dementia services among some minority ethnic populations (19,20). However, the underlying reasons for the differences observed in the present study could not be examined and findings should therefore be interpreted cautiously. Variation in inpatient utilisation was observed across ethnic groups. Further research is needed to understand the factors contributing to these differences. Greater cognitive impairment was associated with higher costs in descriptive analyses, although associations varied between pre- and post-diagnosis multivariable models. This finding is consistent with national and global data demonstrating escalating costs with disease progression (21,22).

Memory clinic services were predominantly accessed pre-diagnosis, consistent with the role of memory services in cognitive assessment and diagnostic pathways (23,24). Post-diagnosis, memory clinic utilisation declined. Previous research has suggested that changes in care pathways following diagnosis and wider disruptions to service delivery during the COVID-19 pandemic may influence patterns of service use (23,24). However, the reasons for reduced utilisation in the present study could not be determined from the available data.

### Limitations

This study has several limitations. First, the analysis was restricted to secondary mental healthcare contacts recorded within SLaM and did not capture primary care, acute hospital care, social care services, medication costs, or informal care. Consequently, the findings reflect specialist mental healthcare utilisation and costs rather than the full healthcare or societal costs associated with MCI and dementia.

Second, the study used routinely collected electronic health record data that were not originally collected for research purposes. As a result, some variables may have been incompletely recorded, leading to potential underestimation of service use. In particular, indirect contacts were recorded at relatively low levels, which may reflect underreporting rather than true patterns of service delivery. Patients living in care homes were eligible for inclusion; however, care home residence was poorly recorded and could not be analysed reliably.

Third, MMSE scores were used as a pragmatic measure of cognitive status because they were routinely available within the dataset. However, MMSE does not provide a comprehensive measure of dementia severity and may be influenced by educational, cultural, and language factors, particularly within ethnically diverse populations. Findings relating to MMSE should therefore be interpreted cautiously.

Finally, the study period spanned 2010 to 2020, including the COVID-19 pandemic, during which substantial changes occurred in diagnostic pathways and service delivery. These changes may have influenced patterns of diagnosis and service utilisation. In addition, findings may not be fully generalisable to regions with different population characteristics, healthcare systems, or service configurations.

## Conclusion

This analysis demonstrated increased NHS mental health service costs among patients who continued to access services following dementia diagnosis, reflecting greater intensity of care. Demographic, clinical, and social characteristics were associated with variation in service costs. Higher inpatient costs were observed among individuals living alone and those with greater cognitive and clinical impairment. Understanding patterns of service utilisation surrounding diagnosis may help inform planning and resource allocation within dementia services.

## Data Availability

The data that support the findings of this study are available from the Clinical Record Interactive Search (CRIS) system at South London and Maudsley NHS Foundation Trust. Restrictions apply to the availability of these data, which were used under licence and ethical approval for the current study and are therefore not publicly available. Researchers may apply for access to CRIS data through the appropriate governance and approval processes.

## Acknowledgements

This research was supported by the National Institute for Health Research (NIHR) Applied Research Collaboration Kent, Surrey, Sussex. The views expressed are those of the authors and not necessarily those of the NHS, the NIHR or the Department of Health and Social Care.

## Author contributions

GE conceptualised the study, conducted the data extraction and statistical analyses, interpreted the findings, and drafted the manuscript.

PM, RP, and CS contributed to the study design, provided methodological and clinical guidance, supported interpretation of the findings, and critically revised the manuscript for important intellectual content.

All authors meet the International Committee for Medical Journal Editors (ICMJE) authorship criteria, contributed to the work described, and read and approved the final manuscript.

## Transparency Declaration

The lead author confirms that this manuscript is an honest, accurate, and transparent account of the work being reported. No important aspects have been omitted, and any discrepancies have been explained.

## Data availability

The data that support the findings of this study were obtained from the Clinical Record Interactive Search (CRIS) system at South London and Maudsley NHS Foundation Trust (SLaM). These data are not publicly available due to patient confidentiality and governance restrictions. Access may be granted to researchers subject to approval by the CRIS oversight committee and appropriate governance procedures.

## Analytic Code Availability

Statistical analyses were conducted using Stata version 15.1 (15)

## Research Material Availability

Not applicable

## Thesis statement

This manuscript is derived from research conducted as part of my doctoral thesis at the University of Greenwich. The thesis is entitled:

*“Cost-effectiveness of non-pharmacological therapies for people living with dementia”*

The submitted manuscript represents a revised and condensed version of one chapter of the thesis. The work has been adapted to meet the journal’s formatting and reporting requirements and has not been published elsewhere in this form.

The thesis will be made publicly available through the University of Greenwich institutional repository. Upon publication of this article, the thesis record will be updated to include a citation and link to the published version of record.

I confirm that I hold the necessary rights to reuse this material and that no third-party copyright restrictions apply.

